# Effectiveness of Financial Incentives for a Pedometer-Based Walking Promotion Program

**DOI:** 10.1101/2022.02.07.22270574

**Authors:** Eriko Komiya, Satsuki Taniuchi, Masatsugu Shiba, Ayumi Shintani, Hiroaki Nakamura

## Abstract

Physical inactivity contributes significantly to poor health and the onset of disease. Physical inactivity is also associated with severe economic burdens. Japan’s Ministry of Health, Labor and Welfare (MHLW) cites the provision of various health promotion incentives aimed at health-indifferent groups, which are made up of individuals less interested in health promotion. This study investigated the relationship between medical costs and a pedometer-based walking program providing monetary incentives based on daily step counts. The study sample included 16,816 citizens aged 40–75 years who lived in Takaishi City and were enrolled in the NIH from October 2016 to March 2018. The results of the ordinal logistic regression analysis showed that participation in a walking promotion program with an HPFI was correlated with a reduction in healthcare costs over 1 year in a sample of Takaishi City residents. In fact, there was a difference of 67,077 yen in the average medical cost per person per year between the walking and control groups. With 1,923 walking group participants, the total medical cost reduction was predicted to be at least 12,898,904 yen. In this study, we evaluated the effectiveness of a walking promotion program with incentives. With program context differing widely from region to region and country to country, future investigations are needed to inform the selection of appropriate incentive schemes for programs offered in other regions and countries.

## Introduction

Physical inactivity contributes significantly to poor health and the onset of disease. According to the World Health Organization, the lack of exercise is the fourth leading cause of mortality risk, and increasing exercise is expected to improve other disorders, such as hypertension, dyslipidemia, and obesity.[1] Physical inactivity is also associated with severe economic burdens.[2] In fact, the authors reported that physical inactivity generates costs of USD 53.8 billion internationally, most of which are paid by the public sector. [2]

Japan’s Ministry of Health, Labor and Welfare (MHLW) cites the provision of various health promotion incentives aimed at *health-indifferent groups*, which are made up of individuals less interested in health promotion.[8] Giles et al. reviewed the literature on the effectiveness of financial incentive interventions in promoting healthy behavior changes among nonclinical adults living in high-income countries.[3] Their results indicated that interventions with financial incentives are more effective in promoting healthy behavior changes compared to usual care or no intervention.[3] Although pedometers have been shown to be effective in promoting activities of daily living in a wide range of age groups, most reported studies were conducted on hospitalized patients, with few including nonclinical adult populations.[4,5,6] Currently, there is insufficient research on the effects of municipal-level programs that distribute pedometers to citizens (including healthy individuals), evaluate their level of activity, and provide incentives according to the amount of walking completed. Therefore, this study investigated the relationship between medical costs and a pedometer-based walking program providing monetary incentives based on daily step counts.

## Materials and Methods

This study used data from a program that provided incentives for daily steps measured by pedometers, implemented from October 2016 to March 2018 in Takaishi City in Osaka Prefecture. The program aimed to promote longevity by encouraging people to start and continue exercising. The program was offered not only to health-conscious individuals but also to those who were either indifferent to health promotion or did not exercise much in their daily lives.

The city’s program was widely publicized through information distributed to all resident homes, the city’s website, and posters around the city. Located in an industrial area of Osaka, the city has a population of 56,000, of which 27% is older than 65 years. Takaishi city residents who wished to participate in the walking program were required to register at the city office, where they received a pedometer (Activity Meter with FeliCa AM-150; Tanita Corp., Tokyo, Japan). Participants could also use the pedometer application on their smartphones.

At their convenience, participants registered their daily step data by bringing their pedometers to special machines installed at community centers, post offices, and stores throughout the city. Depending on the number of steps taken each day, up to 6,000 points were awarded over the course of a year (one point equaling JPY1), which could be exchanged for gift certificates. As there is no private medical insurance in Japan, every resident is required to enroll in one of two public health insurance systems.[6,7] This study used a claims data set from one of these systems, the National Health Insurance (NIH), which covers self-employed people (e.g., farmers and fishermen), retirees and their dependents, and the remaining 25% of the population. This data set included the following information: ID number, the month in which domestic diagnosis codes were assigned in each hospital or clinic, whether claims were inpatient or outpatient, whether the claims were medical or DPC claims (medical claims form), the number of days per month medical treatments were provided, domestic diagnosis codes (mapped to the 10th Revision of the International Statistical Classification of Diseases and Related Health Problems), and the medical fees scale (medical expenses). These data were selected in compliance with rules governing the use of personal information and then prepared for analyses. Each individual was given a temporary ID (standardized for medical cost and pedometer data) that was used to link same individuals during analyses. Individuals with registered step count data were considered program participants, whereas those without such data were not considered participants. The daily step data were used for analyses, and the first month with registered step data was considered the first month of program participation. This research protocol was approved by the Ethics Committee of the Osaka City University School of Medicine.

### Analyses

This study was designed to assess the impacts that the program participation had on healthcare costs. When setting up the control and walking groups, attention needed to be paid to factors other than program participation that could affect medical costs in both groups. In this case, there was no data on factors that might be expected to affect the outcome (i.e., medical history, smoking history, body mass index, etc.). Therefore, to adjust for the background factors affecting healthcare costs of the walking and control groups, we adopted a method to reduce bias as much as possible: determining each participant’s basic monthly medical expenses (baseline medical expenses) at the time of the program participation. Baseline medical expenses were defined as the monthly average of medical expenses for 6 months after the program’s starting point. When setting up the control group, age and baseline medical expenses were divided into 5-year and JPY-5,000 categories, respectively. Nonparticipating individuals with the same data (month and year), gender, age category, and baseline medical expense category as the control group were selected on a one-to-one basis. An ordinal logistic analysis was conducted on medical costs during the first 12 months of participation, with the dependent variable of medical costs and the independent variables of program participation, gender, age, time elapsed since participation, and baseline medical costs. A two-sided 5% significance level was used for all hypothesis tests. R version 4.0.1 (https://www.r-project.org/foundation/) was used for analyses.

## Results

Table 1 shows a summary of the descriptive statistical analyses. The study sample included 16,816 citizens aged 40–75 years who lived in Takaishi City and were enrolled in the NIH from October 2016 to March 2018. The sample had an average age of 64.0 years and was 55% female. The median average cost was 42,865 yen, and 1,923 people (16.6%) participated in the program.

**Table 1.**
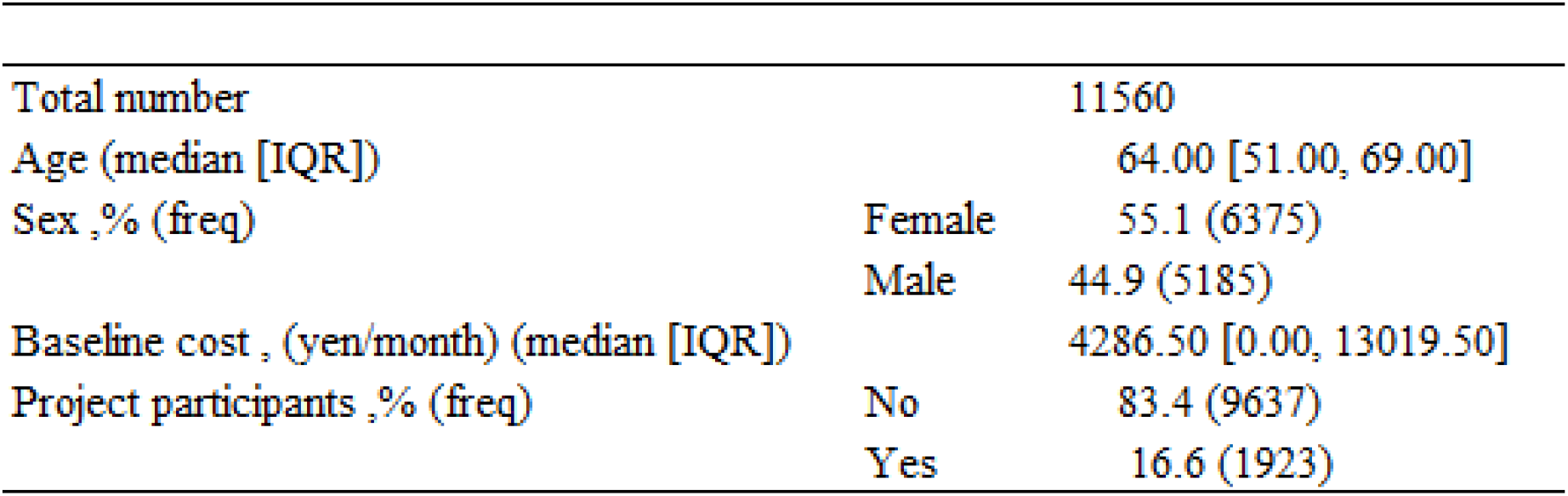
Characteristics of the study samble

One control person with the same background factors was selected for each participant, and the number of subjects analyzed is shown in Table 2. There were no significant demographic differences between the walking and control groups.

**Table 2.** Demographic data for walking group and matched control group

|  |  | Walking | Control | Overall | p Value* |
| --- | --- | --- | --- | --- | --- |
| N |  | 1778 | 1819 | 3597 |  |
| Sex % (freq) | Female | 68.9 (1398) | 68.6 (1421) | 68.8 (2819) | 0.842 |
|  | Male | 31.1 ( 631) | 31.4 ( 649) | 31.2 (1280) |  |
| Age Categories(years old) ,% (freq) | "40-44" | 9.3 ( 189) | 11.1 ( 230) | 10.2 ( 419) | 0.73 |
|  | "45-49" | 9.6 ( 195) | 9.4 ( 195) | 9.5 ( 390) |  |
|  | "50-55" | 8.2 ( 166) | 8.0 ( 166) | 8.1 ( 332) |  |
|  | "55-59" | 8.7 ( 176) | 8.5 ( 176) | 8.6 ( 352) |  |
|  | "60-64" | 12.8 ( 259) | 12.5 ( 259) | 12.6 ( 518) |  |
|  | "65-69" | 25.9 ( 526) | 25.4 ( 526) | 25.7 (1052) |  |
|  | "70-75" | 25.5 ( 518) | 25.0 ( 518) | 25.3 (1036) |  |
| Cost Categories(yen/month) ,% (freq) | "0-4999" | 58.9 (1195) | 59.7 (1235) | 59.3 (2430) | 0.999 |
|  | "5000-9999" | 7.9 ( 161) | 7.8 ( 161) | 7.9 ( 322) |  |
|  | "1000-1499" | 7.4 ( 150) | 7.3 ( 151) | 7.3 ( 301) |  |
|  | "1500-1999" | 5.9 ( 119) | 5.7 ( 119) | 5.8 ( 238) |  |
|  | "20000-2499" | 4.2 ( 85) | 4.1 ( 85) | 4.1 ( 170) |  |
|  | "2500-2999" | 3.4 ( 68) | 3.3 ( 68) | 3.3 ( 136) |  |
|  | "30000-" | 12.4 ( 251) | 12.1 ( 251) | 12.2 ( 502) |  |

\*Comparison between walking and control groups.

Figure 1 shows the program participants’ step registration rate. Results showed that 86.4% of the participants registered their steps for more than 80% of the days in the registration period. Figure 2 and Table 3 shows the results of the ordinal logistic analysis, which indicated that participating in a program with a health-promoting financial incentive (HPFI) was associated with a reduction in healthcare costs over a 12-month period.

**Fig1.**
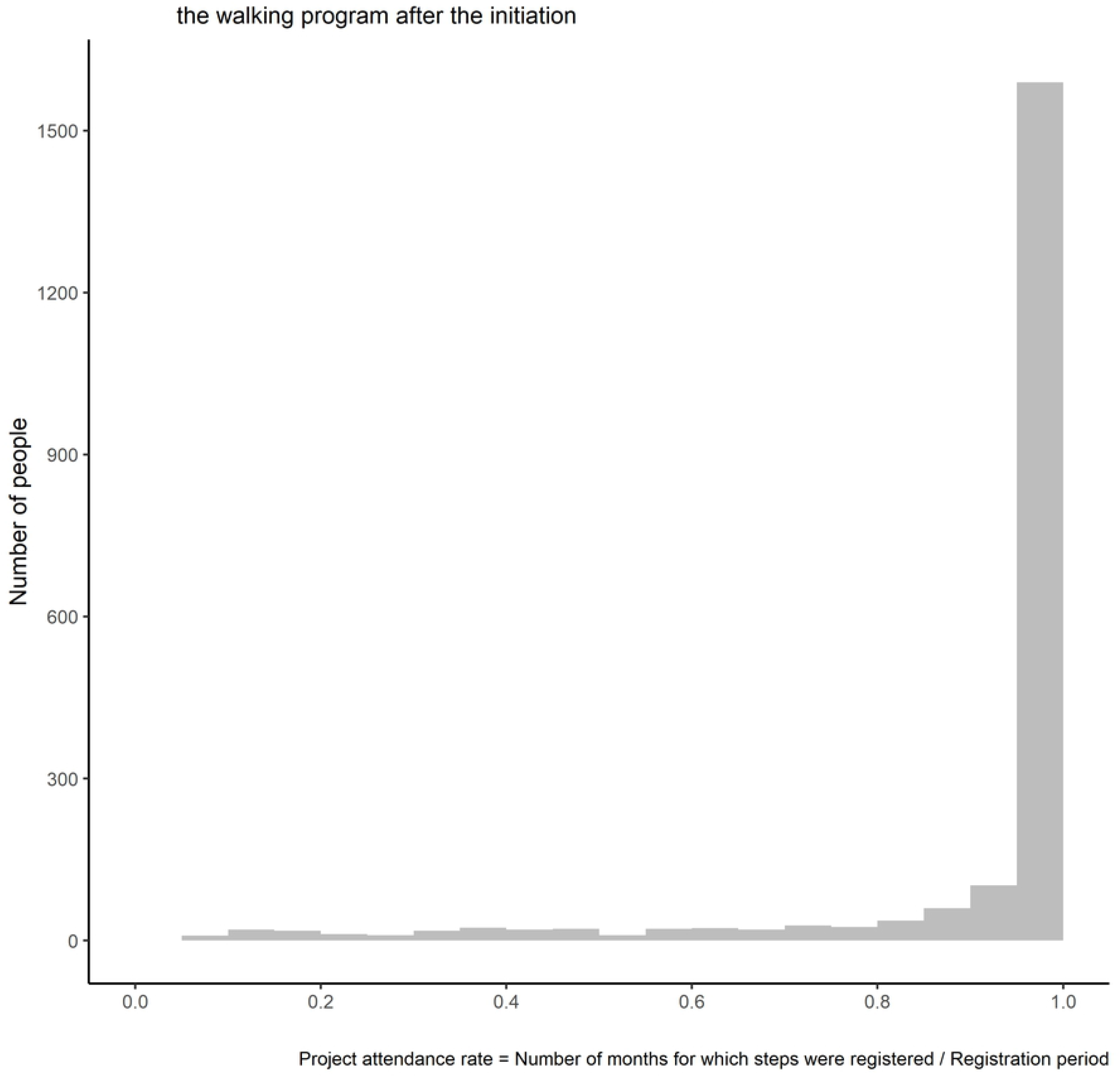
Distribution of percent of months participating

**Fig2.**
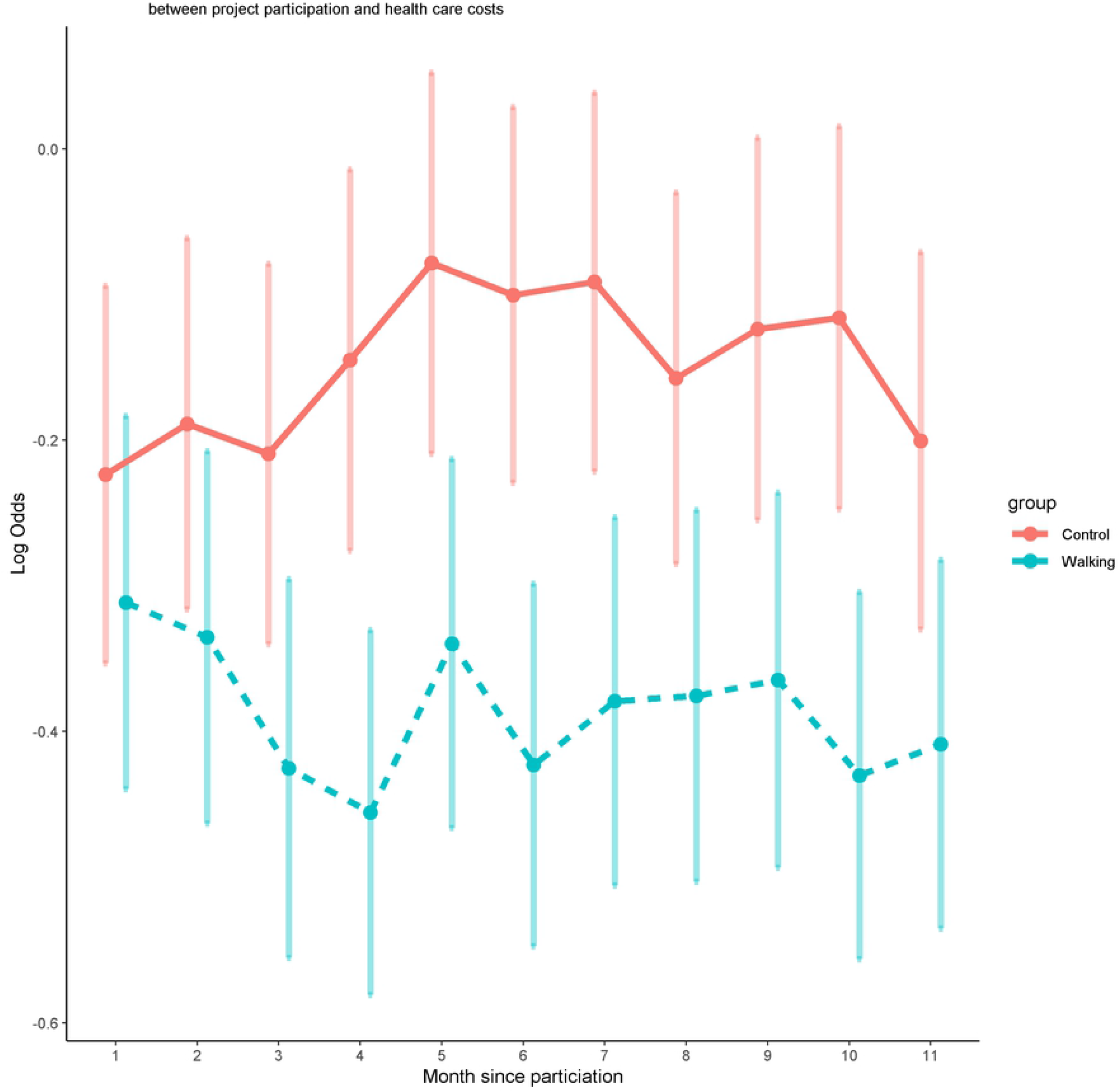
Ordinal logistic analysis plot of the relationship

**Table 3.** Results of ordinal logistic analysis

| Period (month) | Estimate | Lower | Upper | P |
| --- | --- | --- | --- | --- |
| 1 | 0.871 | 0.761 | 0.998 | 0.046 |
| 2 | 0.864 | 0.756 | 0.986 | 0.031 |
| 3 | 0.806 | 0.704 | 0.922 | 0.002 |
| 4 | 0.733 | 0.643 | 0.835 | <0.001 |
| 5 | 0.77 | 0.676 | 0.878 | <0.001 |
| 6 | 0.724 | 0.637 | 0.823 | <0.001 |
| 7 | 0.75 | 0.658 | 0.854 | <0.001 |
| 8 | 0.804 | 0.706 | 0.916 | 0.001 |
| 9 | 0.786 | 0.688 | 0.897 | <0.001 |
| 10 | 0.73 | 0.641 | 0.831 | <0.001 |
| 11 | 0.812 | 0.713 | 0.925 | 0.002 |

The average medical costs per year was JPY 85,525.8 for the walking group and JPY 152,602.8 for the control group (Table 4).

**Table 4.** Average medical costs for 12 months

|  | Walking | Control |
| --- | --- | --- |
| N | 1778 | 1819 |
| Cost average (JPY/year/person) | 85525.8 | 152602.8 |

## Discussion

The results of the ordinal logistic regression analysis showed that participation in a walking promotion program with an HPFI was correlated with a reduction in healthcare costs over 1 year in a sample of Takaishi City residents. In fact, there was a difference of JPY 67,077 in the average medical cost per person per year between the walking and control groups. With 1,923 walking group participants, the total medical cost reduction was predicted to be at least JPY 12,898,904.

With a total of 2,848 program participants, a further reduction in medical costs was predicted for the population aged 20 years or older. Up to 6,000 yen per person per year were given back to citizens according to their daily step count. The average number of points given back was 20,659,200 yen per year, an amount much lower than the total projected medical cost reduction. These results indicate that a municipal program providing incentives of up to 6,000 yen per year based on the daily step count taken via pedometers can help reduce medical costs, with an effect greater than the number of points returned.

Indeed, previous studies have reported that HPFIs may be effective in promoting exercise.[9,10,17–19,21] Finkelstein et al. conducted a two-group, single-blind randomized controlled trial to evaluate the effects of an HPFI that provided monetary incentives according to the number of aerobic exercise minutes recorded by a weekly pedometer over a 4-week period. The results showed that the incentive group had a higher level (1.7 times) of weekly exercise than the control group.[9] Therefore, the researchers concluded that linking a moderate monetary incentive to aerobic exercise minutes is an effective and potentially cost-effective approach to increasing physical activity.[9] Although that report had the advantage of using a randomized controlled design, it had a small sample size of only 51 subjects. In contrast, this study was conducted on a municipal scale and included 1,923 walkers in the analysis, a number larger than previous studies. Furthermore, incentive measures implemented by confirming manual step counts during face-to-face meetings with participants limited the number of people who can participate. To solve this issue, the city of Takaishi operated the program using a system based on information and communication technology. This automated system allowed participants to register their steps by bringing their pedometers to upload sites (registration terminals) set up throughout the city. This approach allowed many participants to be managed with a small number of staff.

The literature includes reports on other examples of HPFIs. Meta-regressions that include a wide variety of health promotion interventions have reported that the intervention effect may decrease with time after the intervention period ends.[9–11] However, statistically significant effects in a smoking cessation program persisted through the 6-month post-intervention follow-up, indicating that the withdrawal of the incentive did not result in a sudden decline in program effectiveness.[20] It has been suggested that HPFIs are more effective in promoting simple, one-time behaviors (e.g., vaccination) rather than complex, sustained behaviors.[12–14] Indeed, there is concern that an HPFI’s effect decreases rapidly after the incentive is withdrawn[15] and that behavior change is less likely to be sustained in the case of one-time HPFIs.[16] These results suggest that although health promotion interventions are associated with short-term behavioral changes, the long-term effects of such interventions require further investigation.

The current walking promotion program with an HPFI was conducted for 2 years and 6 months, from April 2016 to March 2019. Although citizens were given incentives based on their daily step count, there was no penalty for not registering steps. Based on this design, it could be predicted that participants inevitably would register their steps less frequently as their interest in the program waned. As the total participation period depended on the registration start date, this study’s analyses were limited to only 12 months from the registration start date. Of the participant sample, 86.4% registered their daily step count on more than 80% of the days in a year, which is considered a very high participation rate.

This study has some limitations. As the participants voluntarily chose to participate in the intervention, the results may not apply to the general population. Moreover, as the control group did not register a daily step count, it was impossible to determine whether any changes in their step count or medical expenses occurred during the study period.

## Conclusions

These results suggest that a walking promotion program with an HPFI could help reduce healthcare costs. For incentive measures to work effectively, the rewards must be attractive to participants. If the rewards are too small, the incentive will not work. Conversely, the larger the rewards, the higher the program expenses will be as the rewards are paid to participants. This program’s incentives were based on the HPFI guidelines from the MHLW as well as incentives granted by similar policies implemented in Japan. With program context differing widely from region to region and country to country, future investigations are needed to inform the selection of appropriate incentive schemes for programs offered in other regions and countries.

## Data Availability

All datasets are available from the figshare database (https://doi.org/10.6084/m9.figshare.19092023.v1)

## Acknowledgments

We are grateful to Mr M Funatomi for his managing the project site.

